# Genetic polymorphism and risk of anti-tuberculosis drug-induced liver injury (AT-DILI): a systematic review and meta-analysis

**DOI:** 10.1101/2025.01.20.25320827

**Authors:** Amirreza Dehghan Tarazjani, Sima Mohammadi Jouabadi, Soroush Mohammadi Jouabadi, Elnaz Naderi, Miriam Sturkenboom, Fariba Ahmadizar

**Author notes:** Corresponding author: Fariba Ahmadizar; Department of Data Science and Biostatistics, Julius Global Health, University Medical Center Utrecht, Utrecht, the Netherlands Email address Address: heidelberglaan 100, 3584 CX.

## Abstract

Anti-tuberculosis (TB) drugs like isoniazid and rifampin can cause hepatotoxicity leading to treatment termination. Although Genome-wide association studies (GWAS) have identified variants linked to the risk of developing anti-tuberculosis drug-induced liver injury (AT-DILI), findings remain inconsistent. This study aimed to systematically summarise previous observational studies and assess the association between genetic polymorphisms and AT-DILI risk in adults.

We conducted a comprehensive search of PubMed, EMBASE, and Cochrane Libraries until January 10, 2023. The Newcastle-Ottawa Scale checklist was used to assess the methodological quality of the included studies. Pooled odds ratios (OR) with 95% confidence intervals (CIs) were employed using a random-effect model with the I^2^ statistic to estimate the heterogeneity of results.

Our study included 10 studies (n=3,322) of Asian ancestry. We identified genetic variants in drug-metabolizing enzymes, including NAT2, CYP2E1, and PXR, linked to AT-DILI risk. CYP2E1 C1/C1 and slow acetylators of NAT2*6A/6A, NAT26A/7B, NAT27B/7B, and NAT25B/7B genotypes were associated with increased risk, while rapid acetylators of NAT24/4 and NAT24/7B were linked to decreased risk. No significant association was found between CYP2E1B C1/C2, NAT2(*4/*6A, *4/*5B, *5B/*5B) and PXR with AT-DILI risk.

This study revealed that NAT2 slow acetylator genotypes or CYP2E1 C1/C1 are causally linked to AT-DILI risk. The findings suggest that genetic variants in drug-metabolizing enzymes regulated by NAT2 and CYP2E1 are involved in developing drug-induced liver injury in users of anti-TB drugs.

## INTRODUCTION

Tuberculosis (TB), a communicable disease, ranks among the top causes of death from a single infectious disease. It is estimated that over 1.7 billion people have been infected with Bacillus mycobacterium *tuberculosis*. The global incidence of TB peaked around 2003 and has since been declining slowly. According to the World Health Organization (WHO), approximately 10.6 million people were infected with TB in 2021, making a 5% increase from 10.1 million cases in 2020 (1).

For the treatment of pulmonary TB, either the traditional regimen (≥6 months) or a shortened rifapentine-moxifloxacin regimen (four-month) is recommended. The conventional regimen involves an intensive phase of two months followed by a continuation phase of at least four months, using drugs like isoniazid (INH), rifampin (RMP), pyrazinamide, and ethambutol. In contrast, the shortened four-month regimen, consisting of an intensive phase of eight weeks and a continuation phase of nine weeks, employs the same drugs (2). Antituberculosis drugs are associated with a wide range of side effects. The incidence of hepatotoxicity among patients with TB and on anti-tuberculosis drugs is relatively high(3). Anti-tuberculosis drug-induced liver injury (AT-DILI) diminishes treatment effectiveness and can result in treatment failure. Although anti-tuberculosis drugs are not among the primary drugs that cause hepatotoxicity, they play a significant role in incusing it, especially in patients on both isoniazid (INH) and rifampin (RMP), as opposed to single therapy.

Both genetic and non-genetic factors, such as age, sex, race, malnutrition, low body weight, alcoholism, pre-existing liver disease, and chronic hepatitis B and C infections, have been associated with AT-DILI(4). Figure 1 (adapted from Stefan et al., 2011) (5) offers a simplified illustration of Drug-induced liver injury (DILI) mechanisms in TB patients. The figure highlights the different steps in the process, from drug metabolism and bioactivation to hepatocyte damage, immune response, and the clinical manifestations of DILI. DILI is influenced by genetic factors. Variations in key genes involved in drug metabolism, such as N-acetyltransferase 2 (NAT2), cytochrome P450 2E1 (CYP2E1), cytochrome P450 C9 (CYP2C9), and glutathione-S-transferase (GSTM1 and GSTT1 loci), as well as thiopurine S-methyltransferase (TPMT), contribute to the risk of DILI (6).

**Figure 1.**
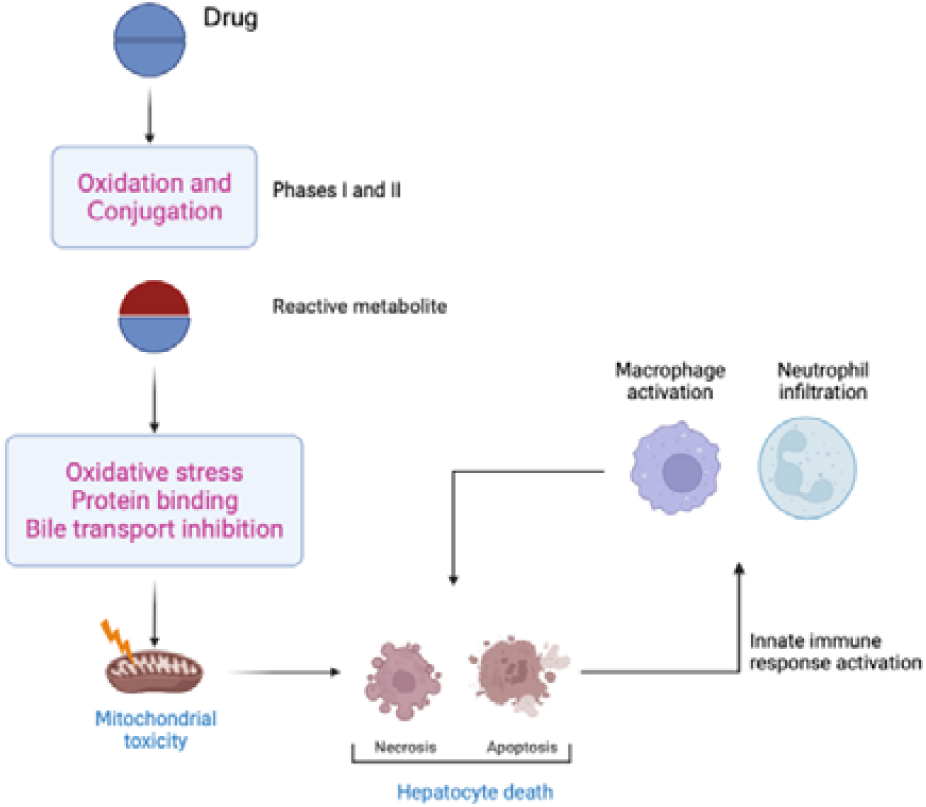
Mechanisms of Drug-induced liver injury Liver P450 systems metabolize drugs in phases I and II. The induction of apoptosis or necrosis by toxic intermediates can result in hepatocyte damage and death. Inflammation can occur when drugs that bind to cellular membranes trigger an immunological reaction upon presentation to major histocompatibility complex (MHC) particles.

Extensive population studies indicate that being a NAT2 slow acetylator elevates the risk of anti-tuberculosis drug-induced liver injury (AT-DILI). Interestingly, smaller-scale genome-wide association studies (GWAS) in European populations did not find a significant NAT2-DILI association. This suggests that race and ancestry could also play pivotal roles in DILI risk and warrant additional investigation(7). Moreover, Emerging evidence points to the involvement of various genes in modulating DILI risk. These include CYP2E1, SOD2, GST isoforms, carboxylesterases (CES) isoforms, and the regulatory factor PXR.(9).

Although existing studies offer valuable insights into the link between genetic factors and AT-DILI, their limitations stem from study design variations and population-specific genetic differences. Consequently, our review seeks to pinpoint the prevalent genetic variants that predispose individuals to AT-DILI. This knowledge will inform personalized prevention strategies for patients taking anti-tuberculosis medications.

## METHOD

The details of the systematic review and meta-analysis were registered in the PROSPERO (registration number: CRD42022300625).

### Search strategy

Following PRISMA guidelines, a systematic literature search of PubMed, Cochrane and EMBASE databases was performed from inception up to January 10, 2023, using the following search string: (’SNPs’ OR ‘single nucleotide polymorphism’ OR ‘Genetic Polymorphism’ OR ‘Genetic variations’ OR ‘Haplotypes’) AND (’AntiTB-induced liver injury,’) AND (’drug-induced liver injury’ OR ‘DILI’). The search was limited to human subjects. A PRISMA flow diagram summarising the study selection process is shown in **Figure 2**. We identified 19 studies that described the relationship between *CYP2E1* or NAT2 or PXR polymorphism and susceptibility to AT-DILI. The study by Vuilleumier, N et al. and the study by Eliana Abreu Santos et al. were excluded due to different ethnicity(8, 10). The study by Dyah A et al. (11). was excluded due to the absence of complete CYP2E1 polymorphism distribution data. The study by Lee, S. W. et al. was also excluded, as it included different treatment protocols and 4 articles discussed other tag SNPs in the *CYP2E1* (12-15). The study by Andrews, E. et al. was excluded due to different populations and ancestors (16). The study by Zazuli et al. was also excluded, as it utilized a different treatment regime (17). Overall, 10 studies were included in the meta-analysis.

**Figure 2.**
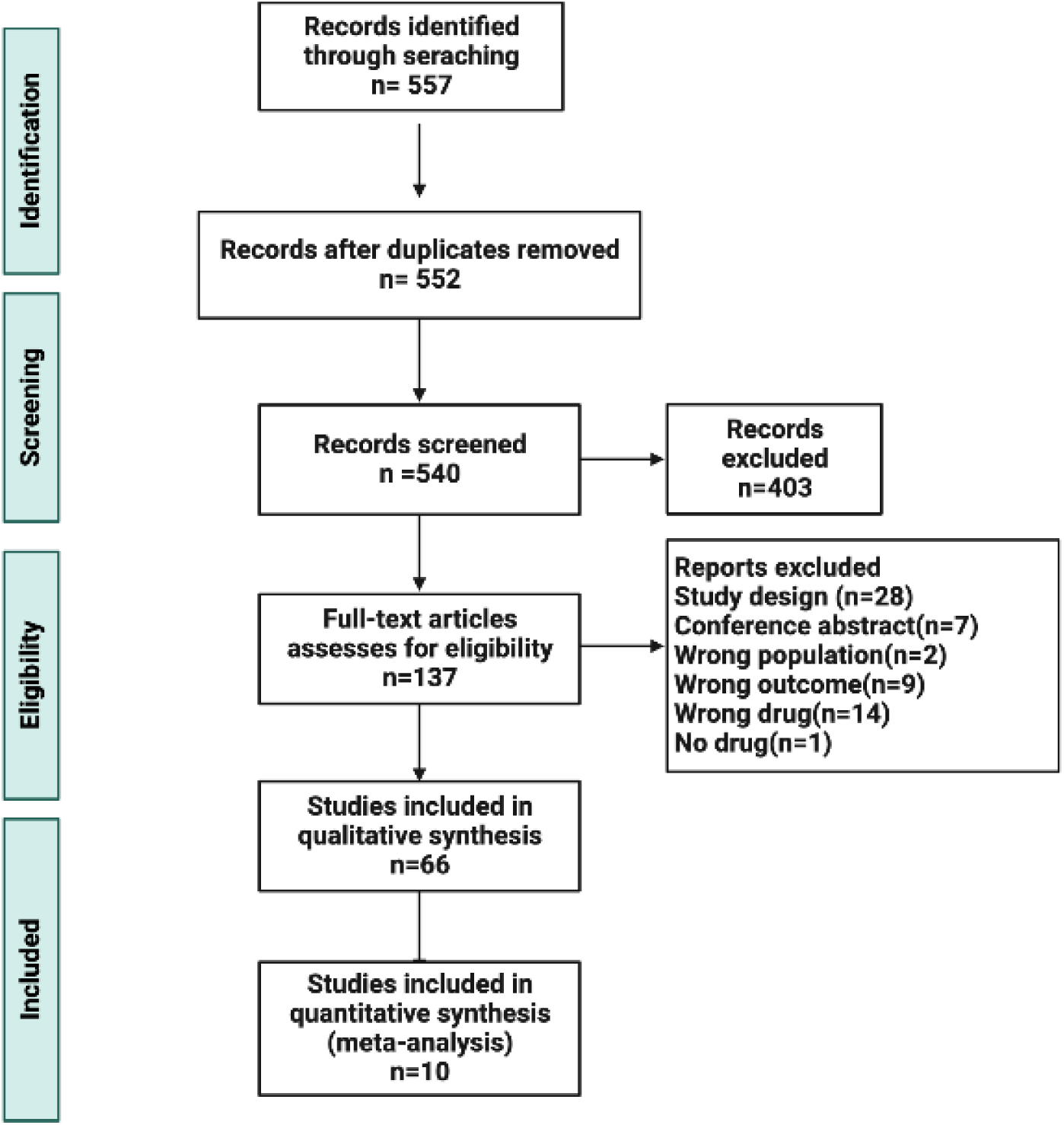
PRISMA flow diagram

The reference lists of the studies included were also manually scanned for potential papers. Two authors independently (AD, SM) screened the retrieved studies according to the inclusion criteria and any discrepancies were solved by the third reviewer’s opinion (FA).

### Inclusion and exclusion criteria

Eligible studies met the following inclusion criteria if they: (i) evaluated the association between single nucleotide polymorphism (SNPs) and risk of DILI in humans (ii) included sufficient data to estimate odds ratios (ORs) and their 95% confidence intervals (CIs).

Studies were excluded if they met the following predetermined criteria: (i) review articles, (ii) studies without complete genetic data for the DILI and non-DILI groups and (iii) animal studies.

### Data extraction and assessment of study quality

The data extracted independently by the two reviewers (AD, SM) included study characteristics such as the name of the first author, publication year, country or region of origin, study type, demographic data of age and gender, setting (clinic), stage of treatment, duration of follow-up, detailed definition of DILI, measurement method for DILI, SNPs, genotyping method and genotype distribution in cases and controls. Study quality was evaluated according to the NEWCASTLE - OTTAWA quality assessment (18) and Reading Mendelian Randomisation studies checklist for GWAS studies (19). Studies with an overall score of ≥5 (range 0 to 10) were considered high quality and retained in the analysis. If any discrepancy occurred, the data were rechecked, and a third author (FA) was invited to give a final decision.

### Statistical analysis

Due to the substantial between-study heterogeneity, we could only perform the meta-analysis on studies with anti-tuberculosis users of Asian ancestry.

We checked whether the OR was calculated for a homogenized approach between alleles. For example, allele C was the reference value in all 10 included studies. We calculated the OR based on A as a comparator versus C as a reference for those studies and vice versa; we recalculated the OR to have a homogenized effect estimate.

Individuals homozygous for rapid *NAT2* acetylator alleles (*NAT2*\**4*) were classified as rapid acetylator phenotype (RAs); individuals who do not carry the NAT2*4 allele or other rapid alleles of NAT2 were classified as slow acetylator phenotypes (SAs) and individuals who have a copy of the *4 and a copy of either *5B, *6A, or *7B, were classified as intermediate acetylator phenotypes (IAs).

We exponentiated beta coefficients to capture the OR from studies where beta coefficients were reported to achieve a homogenised effect estimate. Using a meta-analysis approach, we pooled the calculated and extracted ORs. The meta-analysis was performed using Metan packages of R studio 3.3. Mantel-Haenszel OR was used as the primary statistic to pool the results. Heterogeneity was assessed using I^2^, and random effect models were used when I^2^ was more than 50%. The prediction interval was further calculated for the random effect models to present the range of true effects of included studies. A high I^2^ estimate is not necessarily synonymous with important heterogeneity, so we calculated the p-value to determine the existence of significant heterogeneity.

## Result

### Study characteristics

A total of 557 studies were identified. After deduplication, 540 articles were screened by title and abstract. Out of these, 137 were selected for full-text screening. From this group, 127 studies were excluded due to different study designs, different populations, non-relevant results, different drugs, and studies with no drugs. Eventually,10 studies were finalized for inclusion in the meta-analysis.

Although 66 studies have assessed genetic polymorphism of DILI caused by anti-TB in different races, meta-analysis was not performed in these studies due to the scattering of information and insufficient treatment data (Figure 2).

The main findings of each study included in the meta-analysis and related information are summarised in Table 1, which shows the main characteristics of the 10 studies (9, 20-28).

**Table 1.**
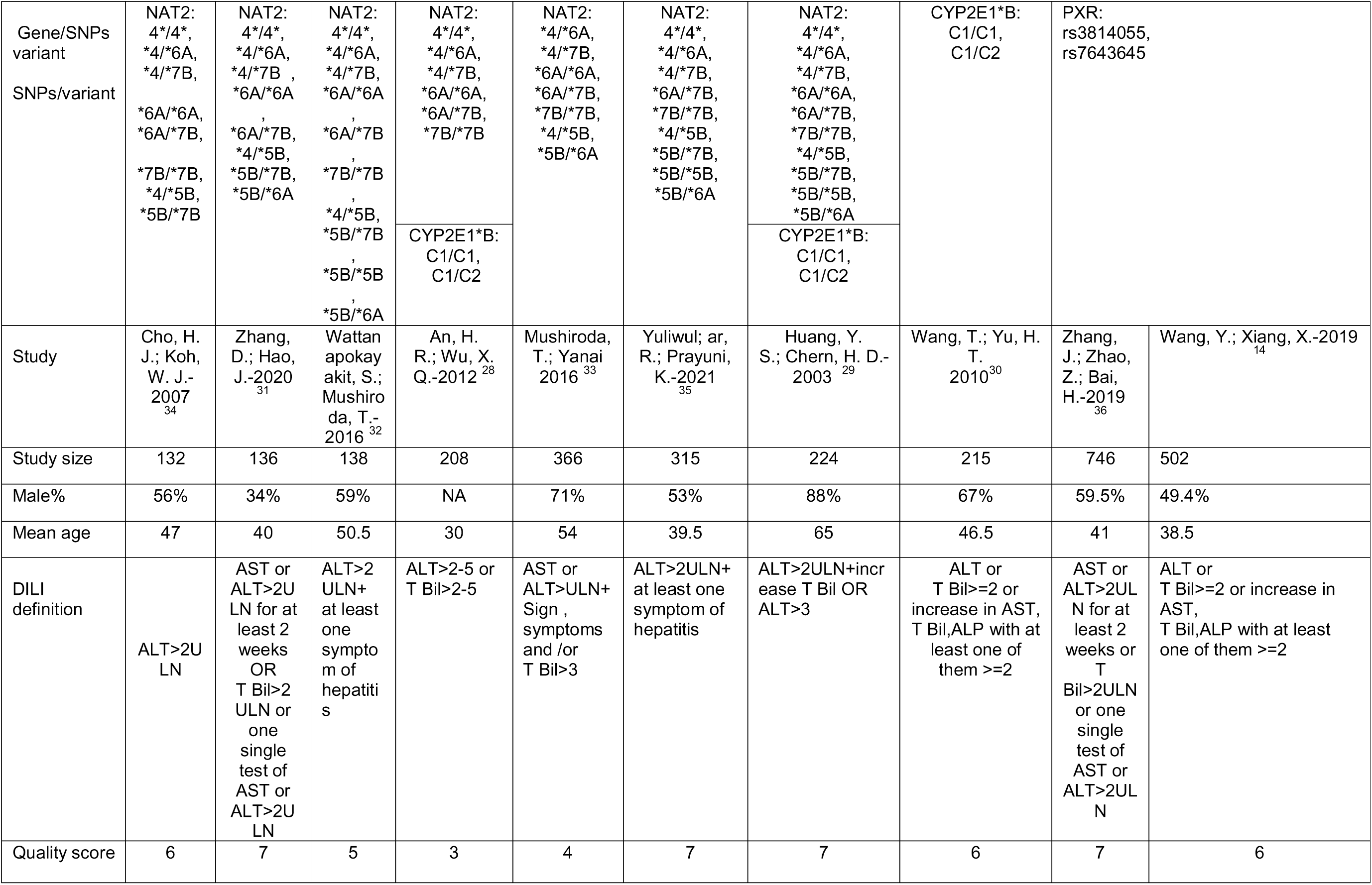
Main characteristics of included studies (n = 10). DILI = drug-induced liver injury; ALT = alanine aminotransferase; AST = aspartate aminotransferase; ULN = upper limit of normal value; T Bil: total bilirubin; N-acetyltransferase 2 = NAT2; cytochrome P450 2E1 = CYP2E1; pregnancy X receptor =PXR

These studies involved 3,322 participants, with sample sizes ranging from 132 to 746. The average age of the participants was 46 years (standard deviation :1.10), and 61% were male. In the case-control studies, “cases” comprised patients with AT-DILI, while “controls” included patients without AT-DILI.

All were candidate gene studies conducted in Asian populations who have received standard anti-tuberculosis regimens. There was no common definition of DILI in these studies. However, the majority of them defined DILI using the criteria of Alanine transaminase (ALT)>2 Upper limit of normal (ULN), but further selection criteria range from Bilirubin (BIL)>2 ULN to at least one symptom of hepatitis.

### Association studies

Table 2 contains information about three important drug-metabolizing enzymes, including NAT2 with 10 major genetic variants, CYP2E1*B and its 2 genetic variants, and PXR with 2 genetic variants.

### N-acetyltransferase and susceptibility to AT-DILI

According to Figure 3, pooled results of 7 studies (n=1,441), NAT2 *4/*4 (RAs)with OR:0.49 (*95%CI 0.36*, 0.66) and NAT2*4/*7B (IAs) with OR:0.40 (*95%CI* 0.27 to 0.58) were associated with a decreased risk of AT-DILI and are significant preventive genetic factors. On the other hand, slow NAT2 acetylators with NAT2*6A/*6A (OR:4.24 [*95%CI 2.87* to 6.26]) and NAT2*6A/*7B (OR:3.78 [*95%CI* 2.11 to 6.76]) were shown as statistically significant associations with increased risk of AT-DILl. In addition, NAT2 *4/*6A (IAs) with OR: 0.82 (*95%CI* 0.53 to 1.24) did not have a significant association with the risk of AT-DILI. When 6 studies (n=1,278) included NAT2*7B/*7B (SAs) and 5 studies(n=867) included NAT2 *5B/*7B (SAs) were pooled in the meta-analysis, a significant association was observed between slow NAT2 acetylators and the risk of AT-DILI. The OR for NAT2*7B/*7B was 2.68 (95%CI 1.30 to 5.51), and for NAT2 5B/*7B was 2.93 (95%CI 1.40 to 6.12).

**Figure 3.**
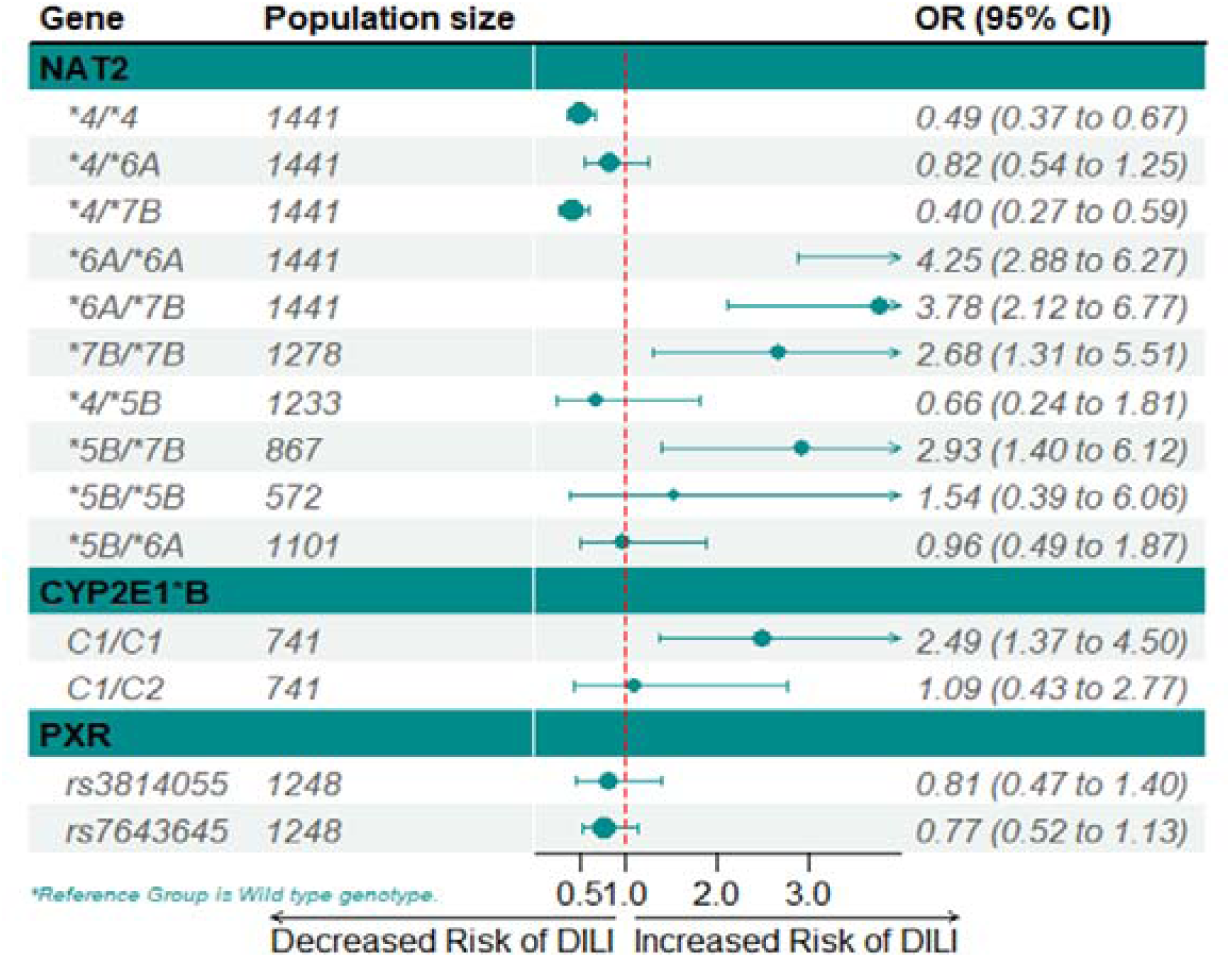
Summarized ORs of DILI associated with haplotypes of NAT2, CYP2E1*B and PXR. ▪ = study-specific ORs (square size reflects the study-statistical weight, i.e., the inverse of the variance); — = 95%CIs. CI = confidence interval; OR = odds ratio; DILI = drug-induced liver injury.

Our meta-analysis showed statistically non-significant associations between NAT2 *4/*5B (IAs), containing 6 studies (n=1,233), NAT2*5B/6A (SAs), having 5 studies (n=1,101) and NAT2 *5B/*5B (SAs), containing 3 studies (n=572) genotypes and susceptibility to AT-DILI (**Supplementary** Figure 1).

### Cytochrome P450 2E1 and susceptibility to AT-DILI

According to Figure 3, the pooled OR of 3 studies (n=741) showed that CYP2E1*B C1/C1 was statistically significantly associated with AT-DILI (OR 2.49 (95% CI: 1.37 to 4.50). A statistically non-significant association existed between CYP2E1*B C1/C2 and AT-DILI risk. (**Supplementary** Figure 2)

### Pregnane X receptor (PXR) and susceptibility to AT-DILI

Pooled results of 2 studies involving 1248 patients (321 in the case group and 927 in the control group) showed a non-significant association between PXR rs3814055 and PXR rs7643645 and risk of AT-DILI with OR of 0.81 [95% CI:0.47 to 1.40) and 0.77 (95% CI: 0.52 to 1.13), respectively (**Supplementary** Figure 3).

The proportion of variability in the meta-analysis of NAT2 4*/*4 was estimated at 18% with a heterogeneity P value of 0.30, which cannot be certainly considered as a homogenous distribution among NAT2 4*/*4 variations; however, I^2^ was estimated at 0% for NAT2 *6A/*6A, *7B/*7B, *5B/*5B, *5B/*6A, CYP2E1*B C1/C1, C1/C2 due to sampling error within studies. NAT2 *4/*6A (heterogeneity P-value: 0.04), *4/*7B (P value: 0.07), *4/*5B (P value: 0.03), *6A/*7B (P value: 0.05), PXR rs3814055(P value: 0.04) had substantial heterogeneity with I^2^ >50. Despite I^2^>50 for NAT2 *5B/*7B and PXR rs7643645, we did not account for heterogeneity due to a non-significant P value.

### Sensitivity analysis

Sensitivity analysis was conducted after eliminating low-quality literature (score <5) to determine whether the literature quality was the source of heterogeneity. The summarised OR ranged from 0,65 95% CI: 0.23, 1.80; when excluding the study by *Zhang et al.* for NAT2 *4/*5B polymorphism; to 0.38(95% CI 0.20-0.74) (23)(Figure 4). For other polymorphisms, changes did not occur for corresponding combined ORs, so our results were statistically robust.

**Figure 4.**
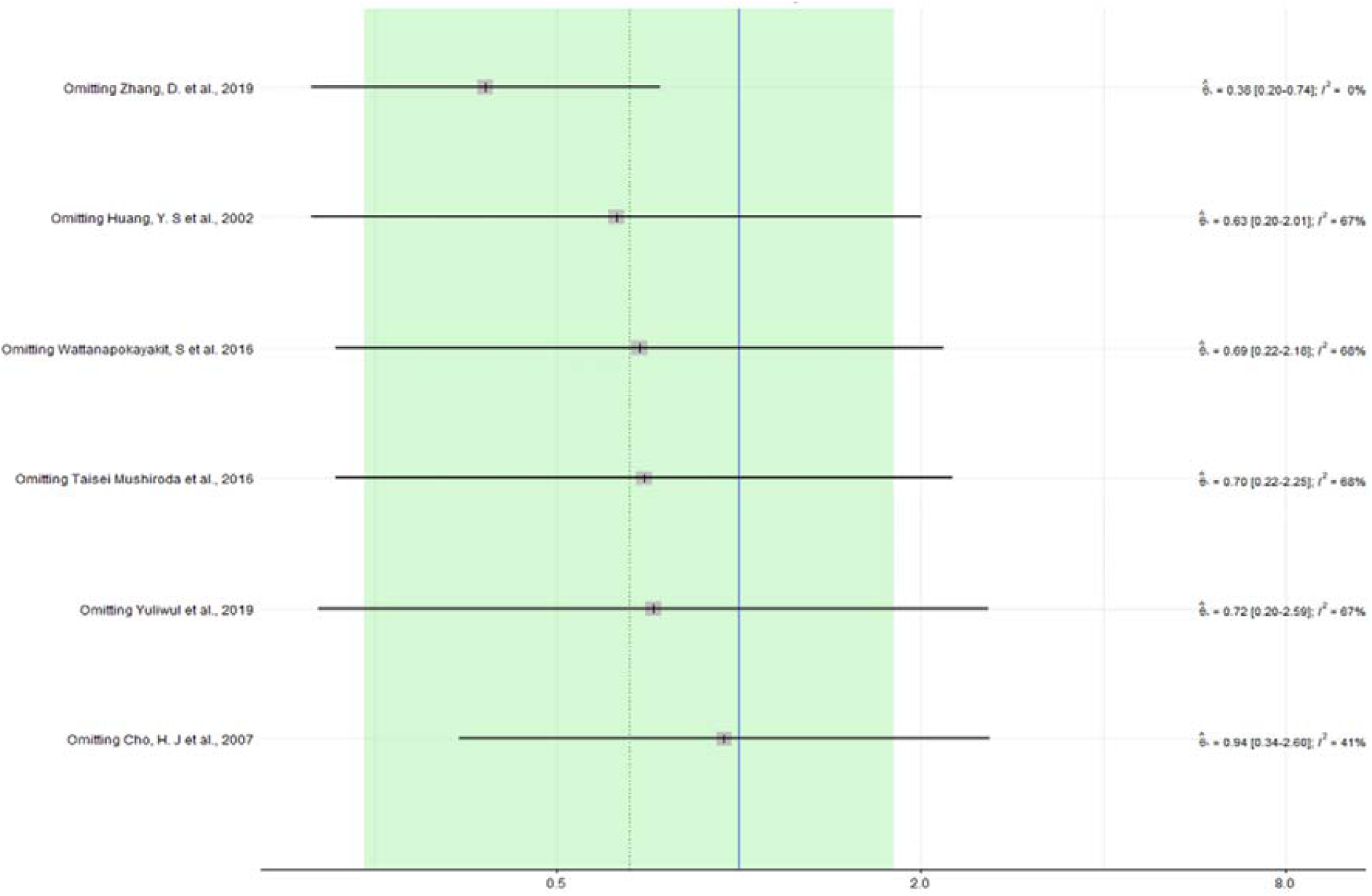
OR ranged from 0,65 (95% CI: 0.23, 1.80) to 0.38 (95% CI 0.20-0.74); when excluding the study by Zhang et al. for NAT2 4/5B polymorphism CI = confidence interval; OR = odds ratio

Egger’s funnel plot asymmetry test in the meta-analysis performed on NAT2 and CYP2E1*B and PXR polymorphism. The study by Wattanapokayakit S et al. had a significant effect on NAT2 *6A/*7B and NAT2 *4/*4, which may stem from the small size of the study compared to other studies(24) (**Supplementary** Figure 4).

## DISCUSSION

### Main finding

In this study, we conducted a comprehensive meta-analysis of 10 studies (n=3,322) highlighting the contribution of genetic variants in drug-metabolizing enzymes, including NAT2, CYP2E1, and PXR, on the increased risk of AT-DILI. Conversely, for the first time, our result showed that both homozygous and heterozygous genotypes of NAT2*4 are associated with a reduced risk of AT-DILI.

Over the past decade, there has been a growing emphasis on personalized dosing therapies, which are tailored based on drug-metabolizing enzymes and transporter genomes. Should the relationship between genetic polymorphisms and AT-DILI risk be established, it would pave the way for creating a personalized clinical drug-dosage model tailored for TB treatment. This model would account for other well-known factors that affect drug exposure (29). Such a personalized model would be particularly beneficial for the populations in South and East Asia, given the high incidence of AT-DILI in these regions. The development of such a model could significantly reduce the occurrence of adverse drug reactions (ADRs) during tuberculosis treatment, particularly in cases where AT-DILI causes treatment interruption. Given the higher cost of treating AT-DILI compared to standard TB treatment, reducing the incidence of ADRs may be cost-effective in countries with a high burden of TB.

### NAT2 and AT-DILI

Genetic variations within the NAT2 gene impact the activity of the NAT2 enzyme. These variations result in distinct acetylator phenotypes: rapid, intermediate, and slow.(30). Rapid NAT2 acetylators carry two copies of NAT2*4, Intermediate acetylators have one NAT2*4 allele and another from the set NAT2*5B, NAT2*6A, or NAT2*7B and Slow acetylators lack the NAT2*4 allele or other rapid NAT2 variants. As illustrated in Figure 5, NAT2, a critical liver enzyme involved in anti-tuberculosis drug metabolism, plays a crucial role. Reduced NAT2 activity can lead to hepatotoxicity due to the accumulation of precursors like hydrazine and acetyldiazene. The efficiency of this detoxification pathway depends on polymorphic alleles. This fundamental detoxification pathway is slower in slow acetylators. Consequently, slow acetylators may increase the concentration of toxic metabolites (31). It has been shown that the NAT2 genotype significantly influences plasma isoniazid levels in Asian TB patients.(32). Past studies have implicated slow acetylation genotypes of NAT2 as genetic factors in AT-DILI(14, 26, 27, 33-35). Our meta-analysis provides compelling evidence that the most significant increase in the risk of AT-DILI was shown in patients with NAT2*6A/*6A (OR= 4.25), NAT2*6A/*7B (OR= 3.78) and NAT2*5B/*7B (OR= 2.93)(20, 21, 36). Careful interpretation of study findings is essential, considering that the significant association between these polymorphisms and the risk of AT-DILI may be influenced by small sample sizes and the low frequency of AT-DILI cases reported in patients with these genetic variants. Some studies have identified a subgroup of slow acetylators within the NAT2 gene, specifically including *NAT2*6A/6A, *NAT2*6A/7B, and *NAT2*7B/7B, which are termed ultra-slow acetylators. Consistent with prior research, our results confirm that these genotypes are linked to a higher DILI risk compared to slow acetylator genotypes. Consequently, patients carrying the NAT2 slow acetylator genotype should receive close monitoring during their tuberculosis treatment.

**Figure 5.**
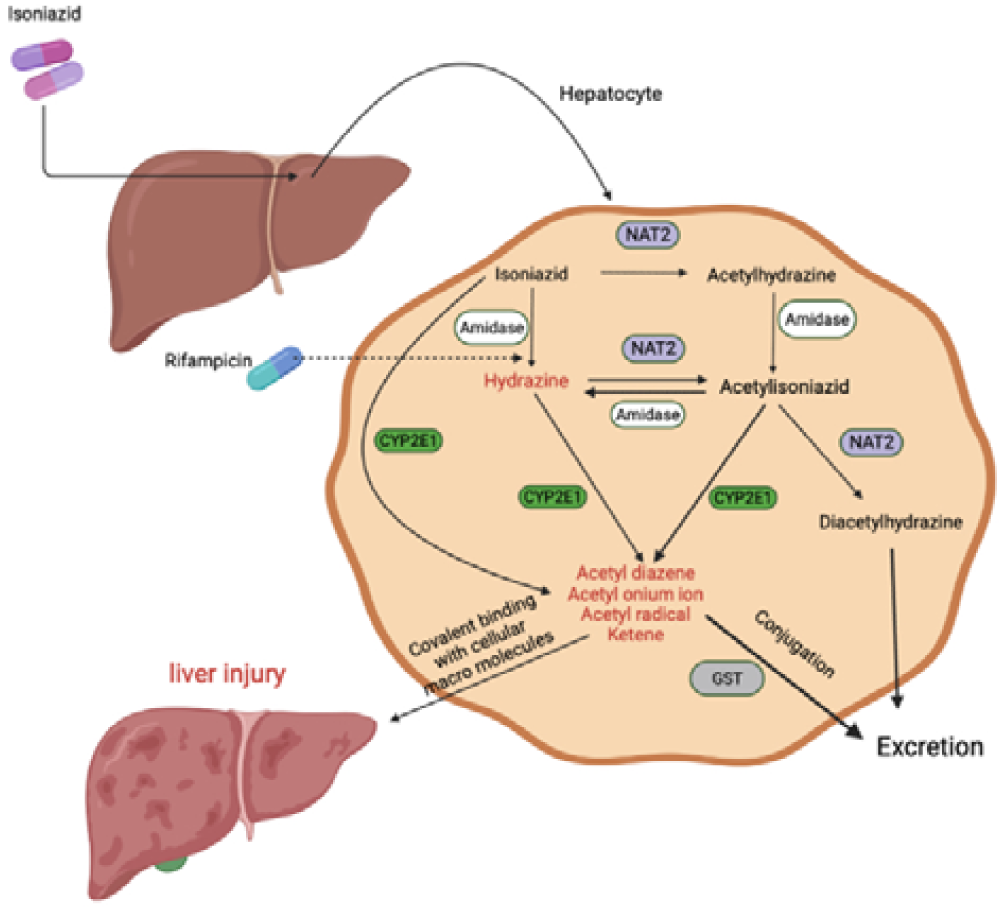
Diagram of interaction between drugs and enzymes that causes anti-tuberculosis drug-induced hepatotoxicity. GST = Glutathione S-transferase; N-acetyltransferase 2 = NAT2; cytochrome P450 2E1 = CYP2E1

**Figure 6.**
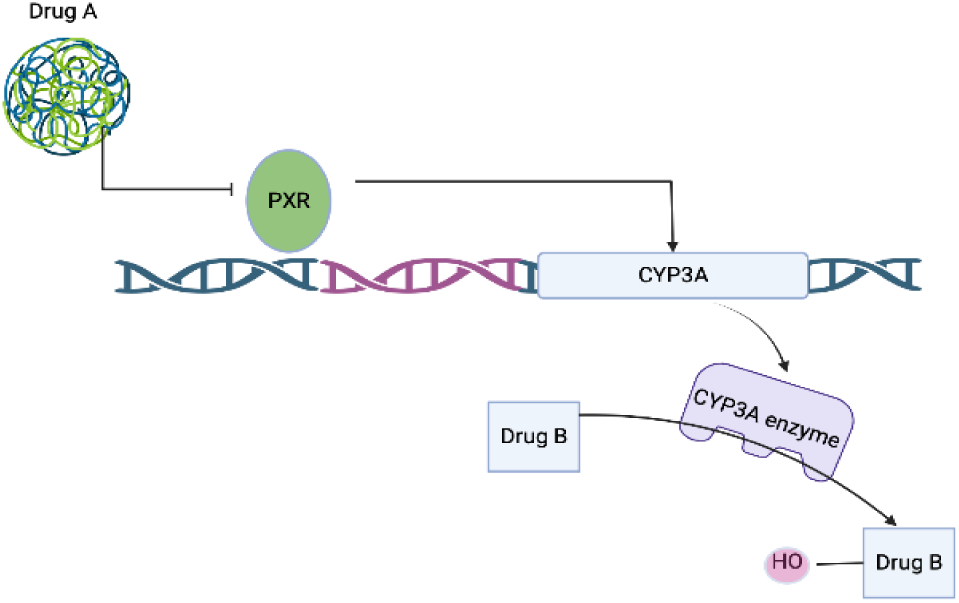
Diagram of interaction between drugs and PXR and enzymes.

Interestingly, our analysis diverges from the study by Mitchell et al., revealing that both homozygous and heterozygous genotypes of NAT2*4 associated with rapid and intermediate acetylator phenotypes can actually mitigate the risk of DILI (37). Among these genotypes, the most significant reduction in AT-DILI risk occurs in patients with *NAT2*4/4.

### CYP2E1 and AT-DILI

We also performed a meta-analysis of 3 studies on the association between polymorphisms in the CYP2E1 gene and DILI. This gene includes three genotypes: C1/C1, C1/C2, and C2/C2, with C1 being the wild allele and C2 as the variant. Consistent with earlier research, our findings suggest a notable rise in the risk of AT-DILI in individuals carrying the CYP2E1 C1/C1 genotype (38-40).

Our data revealed that those with the C1/C1 genotype were 2.48 times more likely to develop AT-DILI. This finding is in line with a previous meta-analysis by Yang S and his team, which reported a 1.39-fold increase in AT-DILI risk in patients with the C1/C1 genotype compared to others(41, 42). In individuals with the C1/C1 variation, the suppressive effect of isoniazid on CYP2E1 was less pronounced than in other patients. Following isoniazid treatment, CYP2E1 activity was found to be elevated in those with the C1/C1 genotype compared to other phenotypes. As a result, individuals with the C1/C1 genotype may produce more hepatotoxins, thereby increasing the likelihood of liver damage(38). The CYP2E1 enzyme is crucial in the metabolic activation of numerous drugs and carcinogens(43). The presence of CYP2E1 can be confirmed using a restriction fragment length polymorphism approach with three restriction enzymes (RsaI, PstI, and DraI). The restriction sites for RsaI and PstI are located in the transcription-regulation region of CYP2E1, which is linked to gene expression(43). A study by Huang et al. (2003) discovered that individuals with the C1/C1 genotype demonstrated higher CYP2E1 activity than those with the C1/C2 genotype during the inhibitory phase of isoniazid(21). Referring to Figure 5, the process of isoniazid metabolism by NAT2 and CYP2E1 involves the direct hydrolysis of isoniazid to produce hydrazine, a harmful metabolite. Acethylhydrazine, the hydrolyzed form of acethylisoniazid, is oxidized by CYP2E1 to produce hepatotoxic intermediates. Additionally, NAT2 acetylation results in a harmless metabolite (diacetyl hydrazine). Moreover, there is a metabolic pathway in which GSTs (GSTM1and GSTT1) detoxify the harmful metabolites produced by NAT2 and CYP2E1(14). A study by Gupta involving 215 individuals from an Asian population found that NAT2 SA combined with CYP2E1 *5 and *6 significantly heightens the risk of DILI incidents(44).

### PXR and AT-DILI

A comprehensive analysis of two studies was conducted to assess the correlation between polymorphisms in the pregnane X receptor (PXR) gene and DILI. PXR, a member of the nuclear receptor superfamily first discovered in 1988, is predominantly expressed in the liver, intestine, and kidney(45). When stimulated by an agonist like rifampicin, PXR enhances the expression of numerous target genes involved in drug metabolism, including the cytochrome P450 family, carboxylesterases, glutathione S-transferases, and UDP-glucuronosyltransferase(46). According to Figure S5, during the combined treatment with multiple anti-TB drugs, PXR could contribute to drug-drug interactions, leading to variability in drug metabolism and disposition(47). Furthermore, PXR activation mediated by rifampicin can serve as a negative regulator of inflammation and immunity, linking drug and xenobiotic metabolism to immune responses by inhibiting the NF-κB signaling pathway (48). In our study, we highlighted the correlation between PXR polymorphisms and DILI risk. We found that the T allele of rs3814055 and rs7643645 in PXR was significantly linked to a reduced risk for AT-DILI, which contrasts with the findings of a study by Zazuli et al. (OR 8.89); 95% CI: 1.36–57.93, P<0.05)(17). This discrepancy could be due to factors such as heterogeneous ethnic origins and varying definitions of AT-DILI. A transition from a C to a T allele was associated with significantly increased transcriptional activity in a study by Manjul Rana (49), suggesting that the rs3814055 C/T polymorphism directly affects PXR’s transcriptional upregulation. In another DILI study, the CC genotype (rs3814055) was linked to an elevated risk of hepatocyte injury due to the decreased expression of CYP3A4, potentially leading to an accumulation of unmetabolized toxic drugs and subsequent hepatocellular injury(16).

It has been proposed that PXR activation by rifampicin could act as a negative modulator of inflammation and immunity by inhibiting the NF-κB signaling pathway (48). Considering the known involvement of the NF-κB pathway in liver injuries and various diseases, it is reasonable to suggest that increased PXR expression could offer a protective mechanism against AT-DILI by suppressing the NF-κB signaling pathway. This potential protective mechanism is consistent with our study’s findings.

### Study Highlights

#### What is the current knowledge on the topic?

Tuberculosis (TB) remains a major global health concern, causing a significant number of infectious disease-related deaths. Despite a decline in global TB incidence, the World Health Organization reported a 5% increase in cases in 2021, reaching 10.6 million. Treatment options, including traditional and shortened regimens, pose challenges due to the associated hepatotoxicity risk.

#### What question did this study address?

This study aimed to investigate the genetic factors influencing Anti-Tuberculosis Drug-Induced Liver Injury (AT-DILI). Specifically, the focus was on the impact of single nucleotide polymorphisms (SNPs) in key drug-metabolizing enzymes - NAT2, CYP2E1, and PXR - on the susceptibility to AT-DILI.

#### What does this study add to our knowledge?

Contrary to previous assumptions, the study reveals that NAT2*4 homozygous and heterozygous genotypes are associated with a reduced risk of AT-DILI. Additionally, comprehensive meta-analyses demonstrate significant associations between NAT2 slow acetylator genotypes, CYP2E1 C1/C1 polymorphism, and increased susceptibility to AT-DILI. Notably, PXR polymorphisms showed a non-significant association with AT-DILI risk.

#### How might this change clinical pharmacology or translational science?

The findings suggest the potential for personalized dosing strategies based on the genetic makeup of individuals, particularly in Asian populations. Identifying NAT2 slow acetylators and those with CYP2E1 C1/C1 genotype may aid in monitoring and implementing preventive measures to minimize the risk of AT-DILI during TB treatment. The study lays the foundation for developing personalized clinical drug-dosage models, contributing to more effective and safer TB treatment.

#### Strengths and limitations

There are several strengths of our study. First, this is the first meta-analysis to evaluate the association between AT-DILI risk and PXR. Second, our study included the most common treatment regime. Third, we included the most updated studies with large sample sizes to better clarify the association of genetic polymorphisms with AT-DILI risk.

Several limitations of this meta-analysis should be mentioned. First, our study was limited to a few studies that met our inclusion criteria. This may have restricted the scope of our analysis and limited the generalizability of our findings. Second, due to the absence of complete distribution data about different ethnicities, our study was limited to the Asian population. Third, due to a lack of information about drug dosing in most studies, we could not account for the effect of anti-tuberculosis drug dosages on AT-DILI risk.

#### Conclusion

Carrying NAT2 slow acetylator or CYP2E1 C1/C1 significantly increased AT-DILI risk during tuberculosis therapy. Clinical monitoring for these patients is strongly recommended. In addition, screening for these genetic polymorphisms may be a clinical advantage to recognizing patients at high risk for AT-DILI and reducing the risk of AT-DILI.

## Data Availability

All data produced in the present study are available upon reasonable request to the authors

## Author Contributions

Conceptualization, F.A.; Methodology, E.N. and F.A and A.D.; Data Curation/analysis, S.M.,Si.M and A.D; Writing – Original Draft Preparation, A.D; Writing – Review & Editing, all authors.

## Supplementary Material Titles

**Figure S1.**
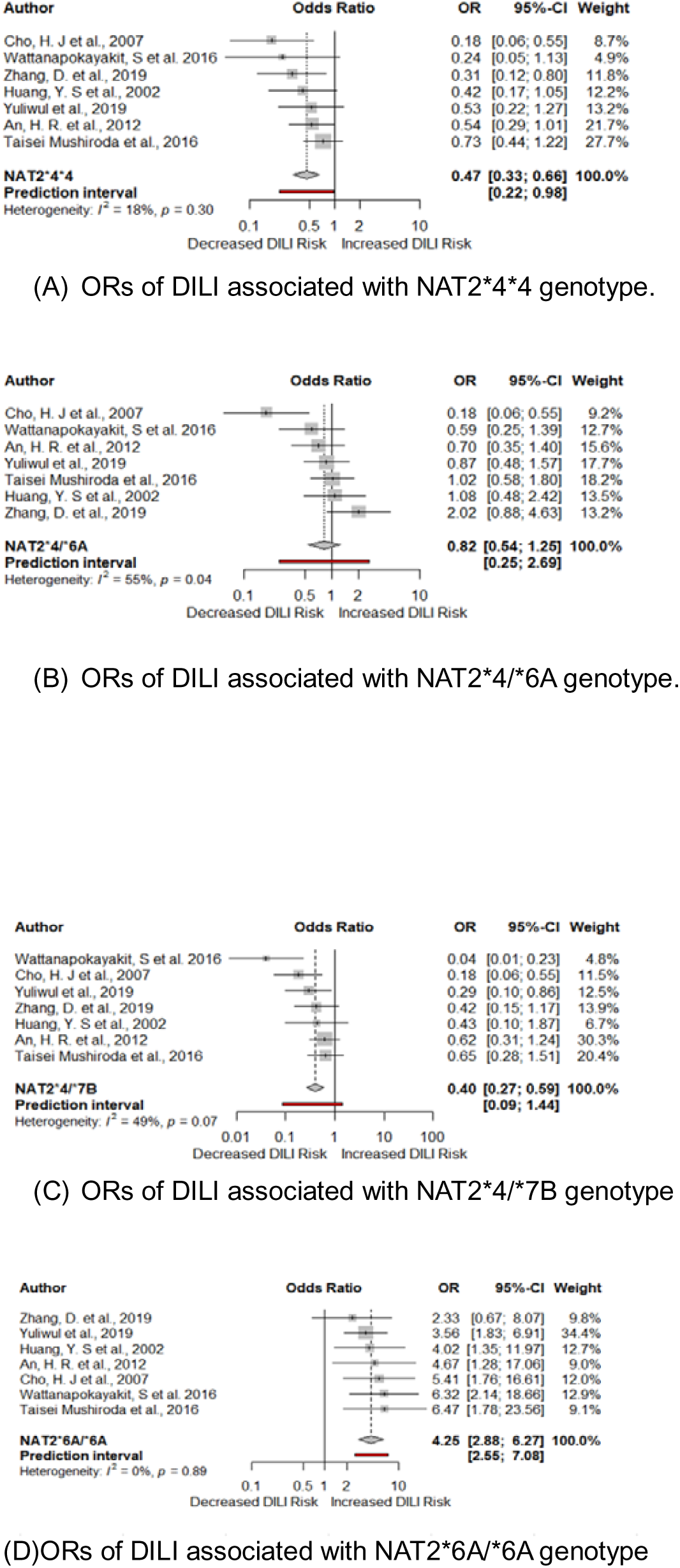
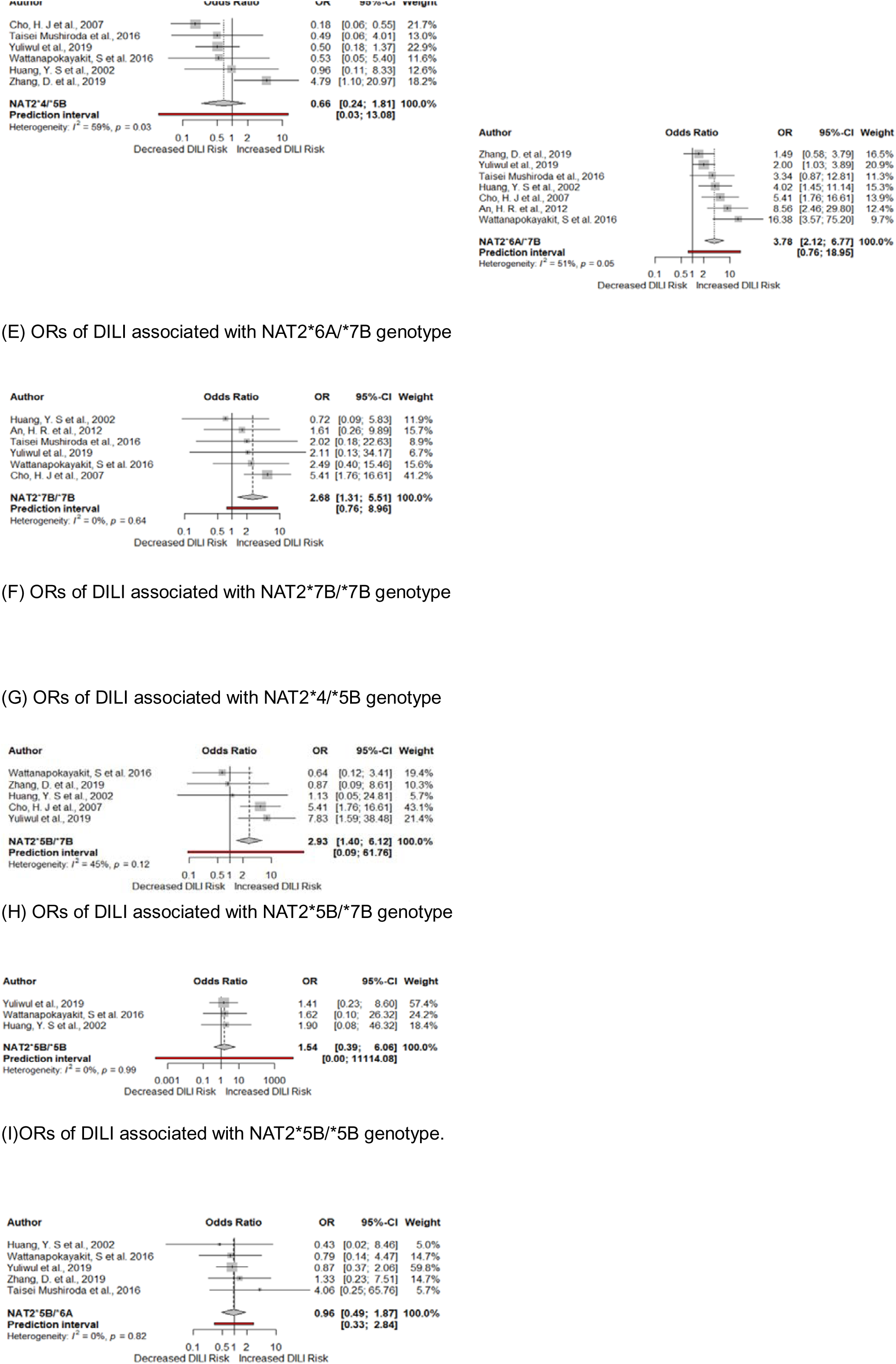
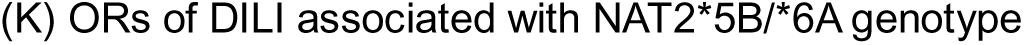
Risk of anti-tuberculosis drug-induced liver injury in patients with the different NAT2 genotypes. ▪ = study-specific ORs (size of square reflects the study-statistical weight, i.e., inverse of variance); — = 95%CIs; ◊ = summary OR with its corresponding 95%CI. CI = confidence interval; OR = odds ratio; DILI = drug-induced liver injury.

**Figure S2.**
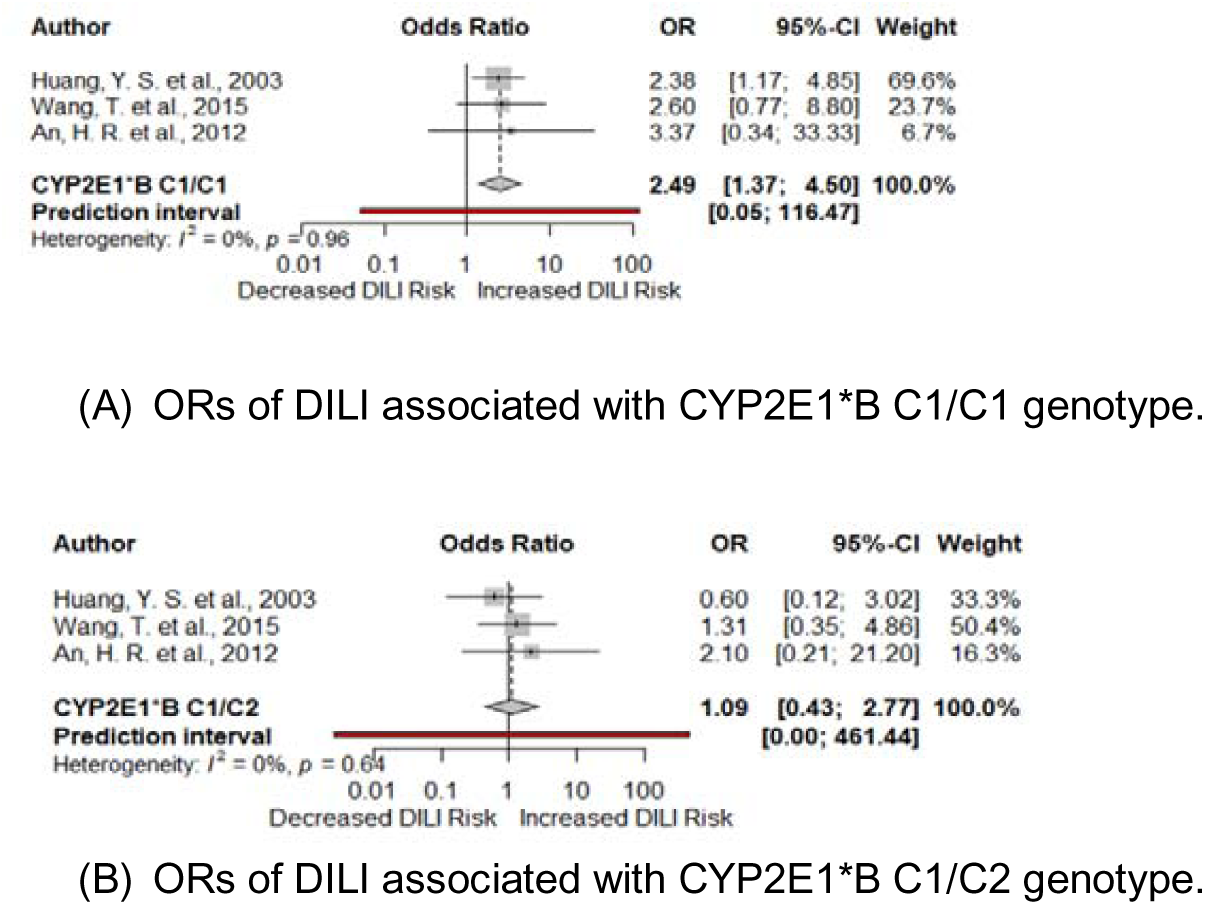
Risk of anti-tuberculosis drug-induced liver injury in patients with the CYP2E1*B C1/C1 and CYP2E1*B C1/C2 genotypes. ▪ = study-specific ORs (size of square reflects the study-statistical weight, i.e., inverse of variance); — = 95%CIs; ◊ = summary OR with its corresponding 95%CI. CI = confidence interval; OR = odds ratio; DILI = drug-induced liver injury.

**Figure S3.**
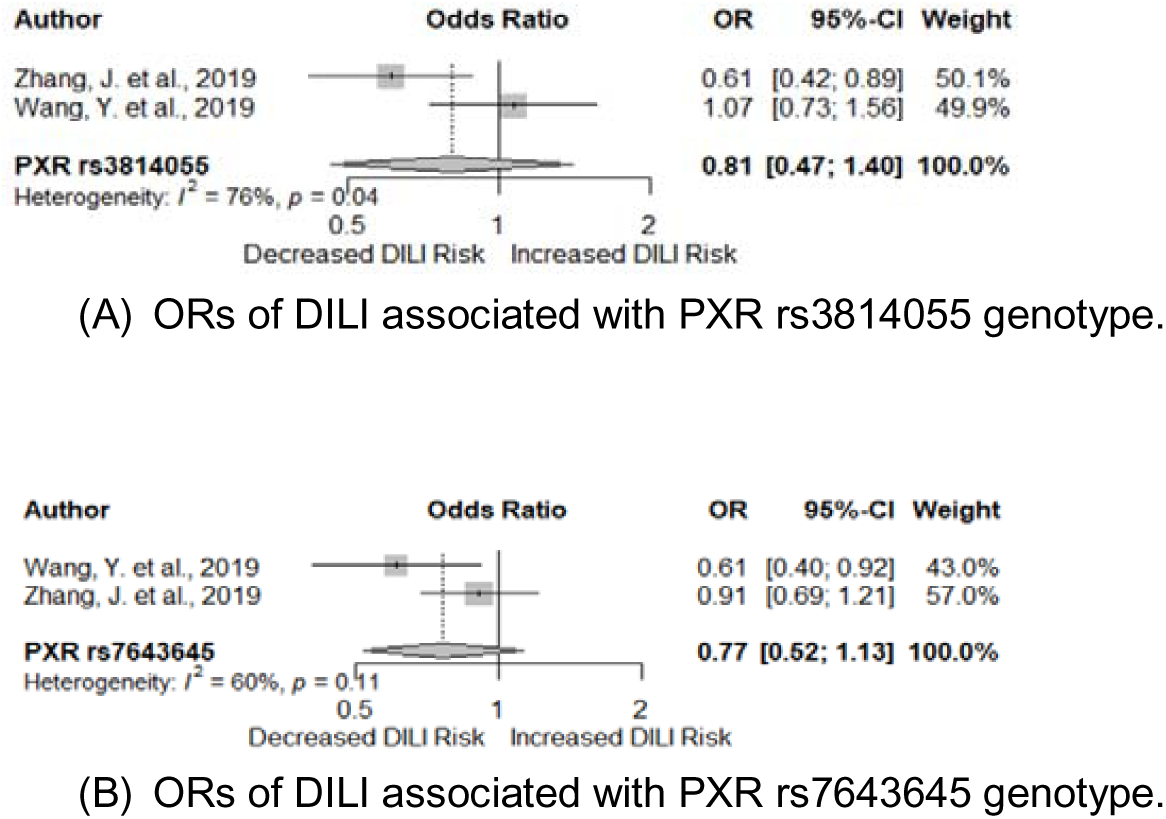
Risk of anti-tuberculosis drug-induced liver injury in patients with the PXR rs7643645 and PXR rs3814055 genotypes. ▪ = study-specific ORs (size of square reflects the study-statistical weight, i.e., inverse of variance); — = 95%CIs; ◊ = summary OR with its corresponding 95%CI. CI = confidence interval; OR = odds ratio; DILI = drug-induced liver injury.

**Figure S4.**
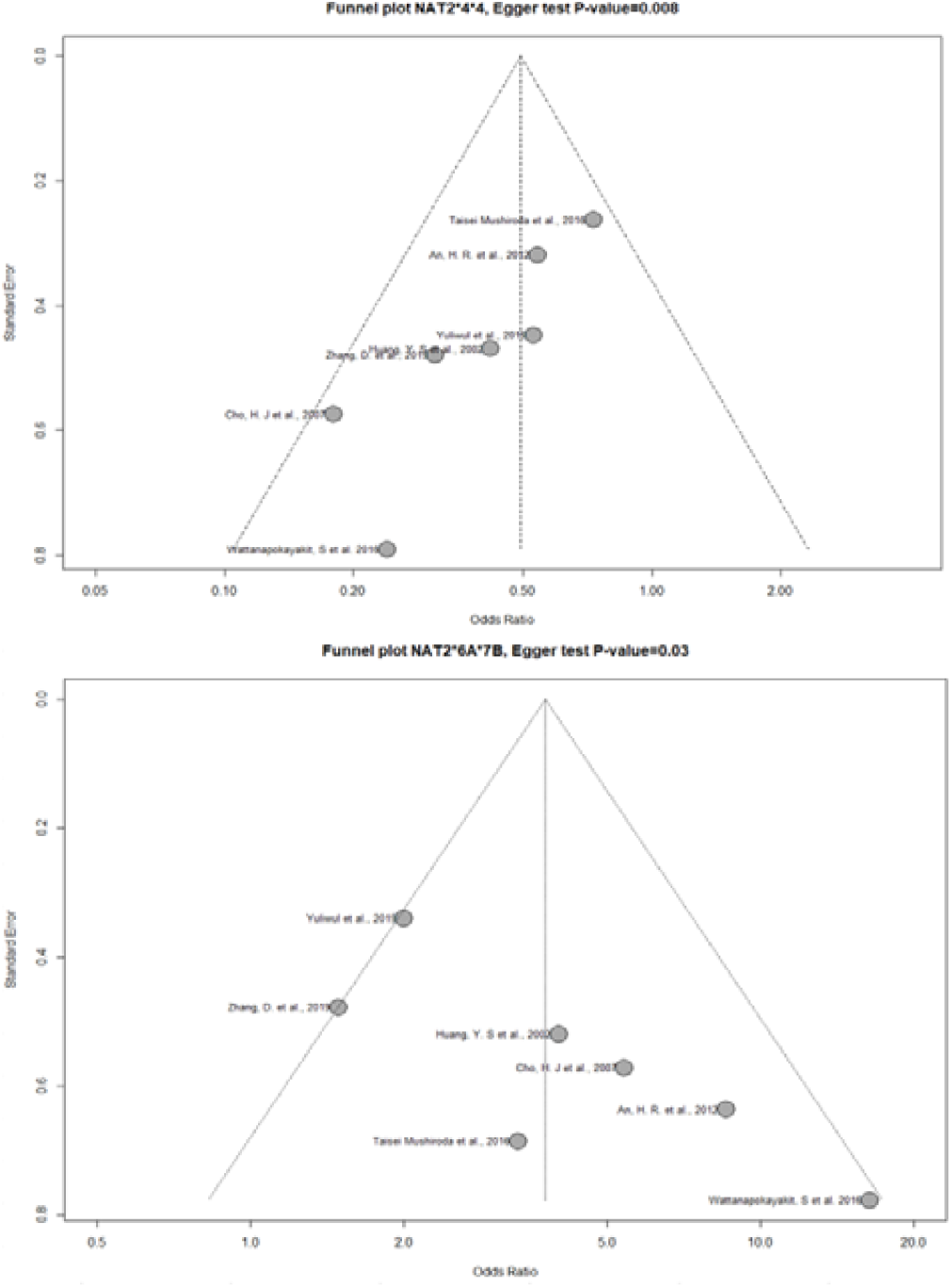
Egger’s funnel plot asymmetry test and affection of the study by Wattanapokayakit on NAT2 *6A/*7B and NAT2 *4/*4.

## legends

**Table 2**. Meta-Analysis of variants associated with AT-DILI

N-acetyltransferase 2 = NAT2; cytochrome P450 2E1 = CYP2E1; pregnancy X receptor =PXR; CI = confidence interval; OR = odds ratio

